# Tuberculosis in Advanced Chronic Kidney Disease

**DOI:** 10.1101/2025.11.26.25341106

**Authors:** Jose Arturo Hernández-Ibarra, Tomas Rafael Mercado-Torres, Jose Raul Ruiz-Ruiz, Carla M Román-Montes, Sandra Rajme-López, Bernardo A Martínez-Guerra, Fernanda González-Lara, Hector O Rivera-Villegas, Zohar Guy, Adriana del C Roblero-Abadía, Alfredo Ponce-de-León, Jose Sifuentes-Osornio, Karla M Tamez-Torres

## Abstract

The management of Tuberculosis (TB) in patients with chronic kidney disease (CKD) presents unique challenges, including an immunosuppressive state, altered drug pharmacokinetics, and limited access to single-drug formulations in our setting. There is a scarcity of real-world evidence on TB outcomes in this population in Latin America. Our study aimed to compare mortality, cure, and relapse rates between TB patients with ACKD and without ACKD.

**Methods:** We conducted an observational, retrospective, age- and sex-matched cohort study that included all patients aged ≥18 years with microbiologically or histologically confirmed TB between 2013 and 2024. Patients with ACKD (GFR <30 mL/min/1.73 m^2^ were compared against age- and sex-matched non-ACKD (GFR ≥30 mL/min/1.73 m^2^) patients. The primary outcome was all-cause mortality at 1 year. Outcomes were compared using the Chi-squared and Mann-Whitney U tests, as well as logistic regression.

**Results:** 51 patients with tuberculosis were included (17 with ACKD, 34 without ACKD). CKD was caused by lupus or diabetes in 29% of patients each. Most CKD patients (68%) received an alternating hemodialysis regimen. One-year all-cause mortality was 18% in both groups (p>0.999), and TB-related mortality was 9% in the control group, vs 0% in the ACKD group. The cure rate was similar between groups (ACKD: 88% vs. non-ACKD: 82%; p=0.586). No relapses occurred. Rheumatologic disease (OR 5.31 (95%CI 1.58-24.38) and hepatotoxicity (OR 20.5 (95%CI 1.82-230.52) were associated with increased mortality, whereas ACKD was not (OR 1.0 (95%CI 0.22-4.61). Due to the low event rate, only bivariate logistic regression was used to identify factors associated with mortality.

**Conclusions:** In our study, ACKD was not associated with one-year all-cause mortality. The alternating hemodialysis-aligned regimen may be a viable strategy in resource-limited settings where individual drug formulations are unavailable. Our findings need to be confirmed in larger prospective studies.

## INTRODUCTION

Globally, around 10.8 million people developed tuberculosis in 2023 (134 per 100,000), with approximately 1.25 million deaths, emphasizing TB’s continued status as a leading cause of infection-related death; the Americas region contributed with approximately 3.2% of incident cases (1,2).

Chronic kidney disease (CKD), including Advanced CKD (ACKD, eGFR <30 mL/min/1.73 m^2^) has been associated with worse outcomes, greater risk for needing renal replacement therapy (RRT), polypharmacy, and a higher risk of infection due to immune dysfunction (3). People with CKD stages 3–5 have an approximately 57% higher adjusted risk of TB than those without CKD, with the most significant excess risk occurring in stages 4-5 (4).

Clinical presentation in CKD patients is often atypical and insidious. Classic systemic symptoms such as anorexia and weight loss can be attributed to those caused by uremia and thus delay the diagnosis of tuberculosis. Additionally, up to 60-80% of these patients present with extrapulmonary disease, including peritoneal tuberculosis, often diagnosed in the context of peritoneal dialysis-related peritonitis (5).

The management of TB in CKD represents significant challenges. Pyrazinamide and ethambutol are renally cleared, thus requiring dose modification. Rifampin has extensive drug interactions and poor penetration to peritoneal fluid, and rifabutin is not available in our region. In CKD patients, individualized non-daily administration of pyrazinamide and ethambutol, careful management of drug interactions, and therapeutic drug monitoring where feasible are recommended (6–9). In our setting, fixed-dose (150 mg rifampicin, 75 mg isoniazid, 400 mg pyrazinamide, and 300 mg ethambutol per tablet) coformulations are universally used. Single formulations of pyrazinamide and ethambutol are either unavailable or scarce, respectively.

While fixed-dose coformulations remain practical, a problem arises when dose adjustments are required based on toxicity, weight, and/or renal or hepatic function.

In our institution, because of the previously addressed limitations, in the last 10 years, it has been frequently used, what we call (and will be hereinafter referred to as) an “alternating hemodialysis dosing”, in which, during the intensive phase, the patient is instructed to take the fixed-dose formulation of 150mg rifampicin/75mg isoniazid/400mg pyrazinamide/300mg ethambutol, 3 or 4 tablets based on weight on the days without hemodialysis, and a fixed-dose 300mg rifampicin/400mg of isoniazid 2 tablets on the days of hemodialysis, after the session (usually three times a week). This would mean that, in the intensive phase, a 70kg adult suffering from ACKD and receiving HD three times per week, would be taking pyrazinamide 23mg/kg 3 times per week, 17mg/kg of ethambutol 3 times per week, rifampicin 600mg daily, and isoniazid 300mg 3 times per week, 800mg 4 times per week.

Evidence from Latin America on clinical outcomes and the effectiveness of pragmatic dosing strategies for TB in patients with ACKD remains scarce. Therefore, this study aimed to describe the rates of all-cause mortality, cure, and relapse in patients with TB with or without ACKD.

## METHODS AND MATERIALS

We conducted an observational, retrospective, age- and sex-matched cohort study at a national tertiary referral center (Mexico City), including adults (≥18 years) with microbiologically or histologically confirmed tuberculosis (2013–2024). We compared patients with advanced chronic kidney disease (ACKD; eGFR <30 mL/min/1.73 m^2^ by CKD-EPI 2021) (2,3) with patients with preserved renal function (eGFR ≥30 mL/min/1.73 m^2^) matched 1:2 (±5 years, same sex). Exclusions: incomplete records and death within 2 weeks of diagnosis. Confirmation required ≥1 of: MTBC culture (Hain or MALDI-TOF), Xpert MTB/RIF or Ultra, tissue IS6110 PCR, or compatible histopathology.

Electronic medical records and clinical charts were reviewed by trained study personnel to gather demographic, microbiological, and clinical information. Patients were followed up for 12 months. Data were accessed on February 2^nd^ 2024. The authors had access to information that could identify individual participants during data collection.

A total of 483 TB cases were identified from 2013 to 2024. We then calculated the eGFR for subjects within ±2 months of tuberculosis diagnosis. Then, we eliminated 181 patients due to insufficient information (n=177) or acute renal disease (n=4), and identified 17 cases with eGFR of <30ml/min. We then randomly selected 34 non-ACKD subjects, matching them by age and sex with the ACKD patients (Fig. 1)

**Figure 1.**
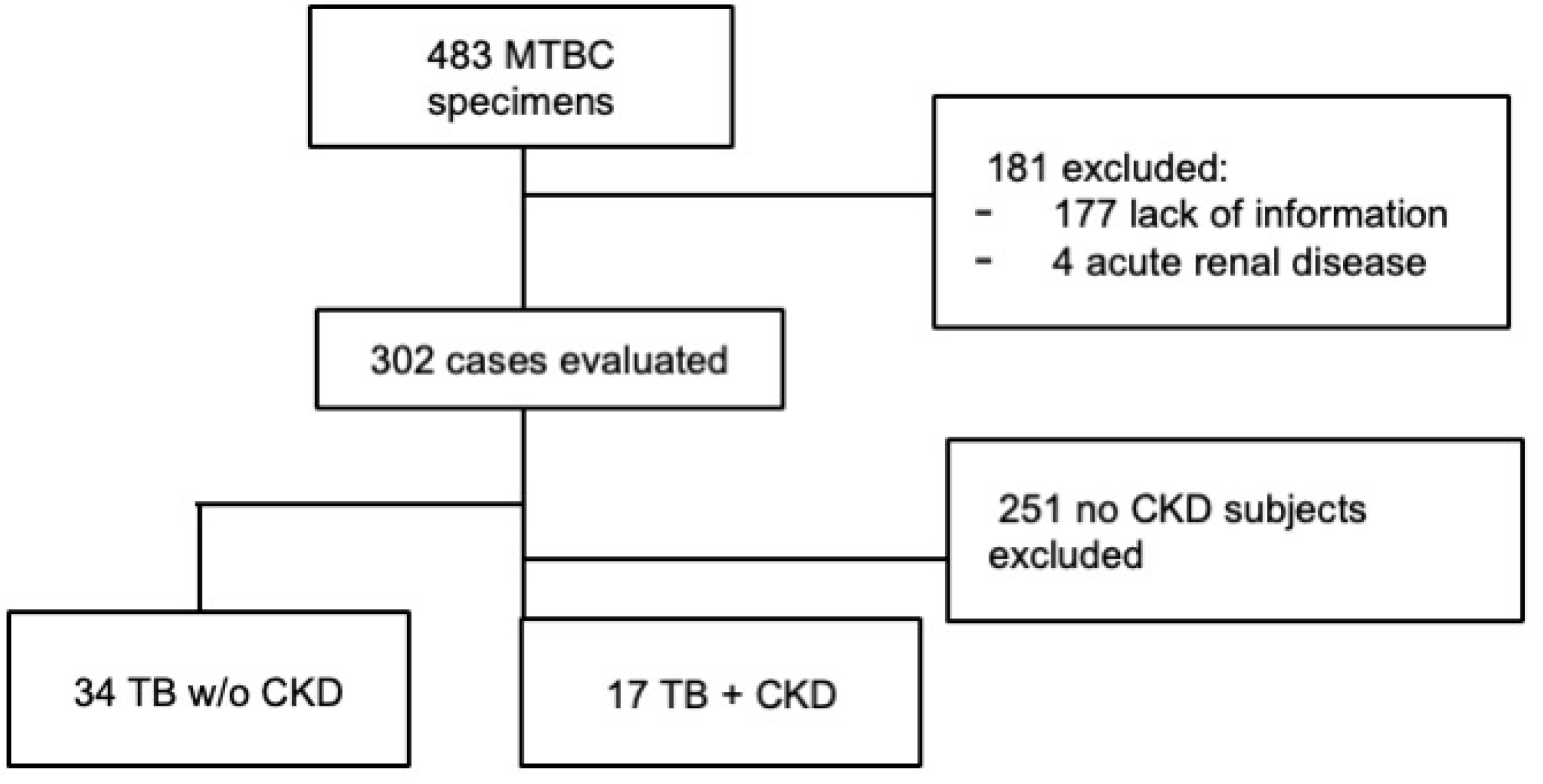
STROBE Flow diagram of the study.

Ethical considerations. Institutional Review Board (Research Ethics and Research Comittees; *Comité de Ética en Investigación y Comité de Investigación*) with local reference number 4922. Informed consent requirement was waived due to the retrospective nature of the study.

### Definitions

Primary outcome was 1-year all-cause mortality; secondary outcomes: TB-attributable mortality, cure, relapse, and adverse events (AEs). Cure and relapse were adjudicated according to standard definitions and were resolved by consensus (8,10). Disseminated TB: ≥2 non-contiguous sites. Hepatotoxicity: ALT ≥3×ULN with symptoms or ≥5×ULN without symptoms (11).

Conventional programmatic regimen used fixed-dose combinations. Where single-agent pyrazinamide/ethambutol were unavailable, an alternating hemodialysis-aligned regimen was used during the intensive phase: DOTBAL® (rifampicin 150 mg/isoniazid 75 mg/pyrazinamide 400 mg/ethambutol 300 mg) on non-dialysis days (3–4 tablets by weight) and DOTBAL-S® (rifampicin 300 mg/isoniazid 400 mg, two tablets) immediately post-hemodialysis (usually three times per week).

### Statistical analysis

Continuous variables are summarized with medians (IQR) and compared using the Mann– Whitney U test, and categorical variables with Fisher’s exact test. Associations with all-cause mortality were estimated using odds ratios (ORs) and 95% confidence intervals (CIs) in bivariable logistic regression. Multivariable modeling was not performed due to the limited number of events (n=9), two-sided α=0.05. Analyses used Stata v14. (StataCorp LLC, Texas, USA).

#### Ethics

This study was approved by the Institutional Review Boards (reference INF-4922-24-24-1).

## RESULTS

Median age 42.4 years (IQR 26–61); 37% were male. Diabetes was documented in 35% (6/17) of ACKD patients compared to 18% (6/34) of non-ACKD patients (p=0.161). HIV was present only in the control group (21%, 7/34, p = 0.044), and rheumatologic diseases were reported in 29% (5/17) of ACKD versus 24% (8/34) of non-ACKD (p = 0.650). A total of 17 patients met the criteria for ACKD and TB and were compared to 34 TB patients without ACKD. Baseline characteristics and comorbidities are detailed and contrasted in Table 1.

**Table 1.** Demographic characteristics and main comorbidities between ACKD and non-ACKD patients.

| Characteristic | Total (N=51) | TB +ACKD<br>(n=17) | TB + non-ACKD<br>(n=34) | p-value |
| --- | --- | --- | --- | --- |
| **Age, years - Median (IQR)** | 42.4 (26-61) | 43.8 (31-61) | 41.8 (25-60) | 0.596 |
| **Male sex - n (%)** | 19 (37.3) | 7 (41.2) | 12 (35.3) | 0.682 |
| **Type 2 diabetes (T2D) - n (%)** | 12 (23.5) | 6 (35.3) | 6 (17.6) | 0.161 |
| **HIV - n (%)** | 7 (13.7) | 0 (0) | 7 (20.6) | 0.044 |
| **Heart failure (HF) - n (%)** | 7 (13.7) | 3 (17.6) | 4 (11.8) | 0.565 |
| **Rheumatologic disease - n (%)** | 14 (27.5) | 5 (29.4) | 9 (26.5) | 0.824 |
| **Any-cause immunosuppression** | 17 (33.3) | 6 (35.3) | 11 (32.3) | 0.834 |

Abbreviations: ACKD, advanced chronic kidney disease; IQR, interquartile range; HIV, Human immunodeficiency virus.

Leading etiologies of ACKD were lupus nephritis with 29% (5/17) and diabetic kidney disease, also with 29% (5/17). Most patients received renal replacement therapy (RRT) (94%, 16/17): hemodialysis (HD) in 82% (14/17) and peritoneal dialysis (PD) in 12% (2/17). 24% (4/17) of patients had a prior kidney transplant with a failed allograft.

The lung was the most frequent site of TB in our study; 75% (12/17) of ACKD patients had pulmonary involvement, with the remainder having exclusively extrapulmonary disease. Disseminated TB was observed in 47% (8/17) of ACKD patients and 74% (25/34) of the non-ACKD group.

Outcomes, disease patterns, microbiology, drug regimens, and adverse effects are shown in Table 2. Cure was documented in 88% (15/17) of the ACKD group vs. 82% (28/34) of the non-ACKD group (p=0.586), and relapse occurred in 0% vs 5.9% (2/34) (p=0.308). One-year all-cause mortality was 17.6% (3/17, 6/34) in both groups (p>0.99), and TB-attributable mortality was 0% vs 9% (3/34) (p=0.20).

**Table 2.** Outcomes, microbiological data, treatment regimens, and outcomes.

| Variable | Total (N=51) | TB+ ACKD<br>(n=17) | TB + non-ACKD<br>(n=34) | p-value |
| --- | --- | --- | --- | --- |
| **Cure - n (%)** | 43 (84.3) | 15 (88.2) | 28 (82.4) | 0.586 |
| **Relapse - n (%)** | 2 (3.9) | 0 (0) | 2 (5.9) | 0.308 |
| **All-cause mortality - n (%)** | 9 (17.6) | 3 (17.6) | 6 (17.6) | >0.99 |
| **TB-attributable mortality - n (%)** | 3 (5.9) | 0 (0) | 3 (8.8) | 0.220 |
| **Disseminated TB - n (%)** | 33 (64.7) | 8 (47.1) | 25 (73.5) | 0.062 |
| **Positive initial culture - n/N (%)** | 47 (92.2) | 15 (88.2) | 32 (94.1) | 0.461 |
| **Positive acid-fast bacilli smear - n/N (%)** | 11 (21.6) | 3 (17.7) | 8 (23.5) | 0.630 |
| ***Mycobacterium* species - n/N (%)** |  |  |  |  |
| *M. bovis* | 21/47 (44.7) | 6/15 (40.0) | 15/32 (46.9) | 0.659 |
| *M. tuberculosis* | 26/47 (55.3) | 9/15 (60.0) | 17/32 (53.1) | 0.659 |
| **Drug resistance - n/N (%)** | 6/47 (12.8) | 2/15 (13.3) | 4/32 (12.5) | 0.936 |
| **Treatment regimen - n/N (%)** |  |  |  |  |
| **Conventional** | 25 (49) | 2 (12) | 23 (67.6) | <0.001 |
| **Hemodialysis-alternating** | 12 (23.5) | 12 (70.6) | 0 (0) | <0.001 |
| **Moxifloxacin addition** | 6 (11.8) | 1 (5.9) | 5 (14.7) | 0.357 |
| **Hepatotoxicity - n (%)** | 4 (7.8) | 1 (5.9) | 3 (8.8) | 0.713 |
| **Other adverse events - n (%)** | 9 (17.6) | 2 (11.8) | 7 (20.6) | 0.699 |

**ACKD, advanced chronic kidney disease.**

Disseminated TB occurred in 47% (ACKD) vs 74% (non-ACKD) (p=0.062). Initial culture positivity was 88% in both groups (47/51); smear positivity was 12% (2/17) vs 27% (9/34) (p = 0.229). Among the 47 cases with species identification, *M. bovis* accounted for 40% (6/15) in the ACKD-group vs 47% (15/32) in the control group (p=0.659), and *M. tuberculosis* accounted for 60% (9/15) vs 53% (17/32) (p=0.659). Resistance to any antitubercular drug (excluding those *M. bovis* strains resistant to pyrazinamide) was observed in 13% of both groups (2/15, 4/32). Among ACKD patients, the alternating regimen was indicated in 71% (12/17). None of the non-ACKD had this regimen indicated. Of the remaining ACKD patients (5/17), two were treated with conventional treatment without dose adjustments, and three (two of whom were on PD) were treated with three daily tablets of DOTBAL (instead of 4). The other two patients in PD were switched to HD and received the alternating regimen.

Hepatotoxicity occurred in 5.9% (1/17) vs 8.8% (3/34) (p=0.713); the case in the first group led to a temporary suspension of treatment. Other adverse effects, primarily gastrointestinal, were reported in 12% (2/17) of patients vs 21% (7/34) (p=0.699). Grading of events was not performed. Resistance to INH and RIF was observed in two cases, and to streptomycin in the other four.

In the bivariate analyses (Table 3), rheumatologic disease (56% vs 19%; OR, 5.31; p = 0.032) and hepatotoxicity (33% vs 2.4%; OR, 20.5; p = 0.014) were associated with all-cause death. ACKD prevalence was identical among deceased and survivors (33%, [3/9] vs 33%, [14/42]; OR 1.0; >0.99).

**Table 3.** Bivariate analysis of factors associated with all-cause mortality.

| **Variable** | **Survivors**<br>**(n=42)** | **Deceased**<br>**(n=9)** | **OR (95%CI)** | **p-value** |
| --- | --- | --- | --- | --- |
| ACKD - n (%) | 14 (33.3) | 3 (33.3) | 1.0 (0.22-4.61) | >0.99 |
| Age, years - Median (IQR) | 41.5 (30-61) | 29 (22-55) | 0.98 (0.94-1.02) | 0.387 |
| Male sex - n (%) | 16 (38.1) | 3 (33.3) | 0.81 (0.18-3.71) | 0.789 |
| Type 2 diabetes | 10 (23.8) | 2 (22.2) | 0.91 (0.16-5.13) | 0.919 |
| Heart failure | 5 (11.9) | 2 (22.2) | 2.11 (0.34-13.15) | 0.422 |
| Rheumatologic disease - n (%) | 9 (21.4) | 5 (55.6) | 4.58 (1.01-20.69) | 0.048 |
| Any-cause immunosuppression | 7 (77.8) | 10 (23.8) | 11.2 (1.99-62.82) | 0.006 |
| HIV infection | 6 (14.3) | 1 (11.1) | 0.75 (0.08-7.13) | 0.802 |
| Hepatotoxicity - n (%) | 1 (2.4) | 3 (33.3) | 20.5 (1.82-230.52) | 0.014 |
| Other adverse events | 8 (19.1) | 0 | - | 0.154 |
| Disseminated TB - n (%) | 25 (59.5) | 8 (88.9) | 5.44 (0.62-47.56) | 0.126 |
| Conventional regimen | 21 (50) | 4 (44.4) | 0.80 (0.19-3.40) | 0.763 |
| Alternating regimen | 11 (26.2) | 1 (11.1) | 0.35 (0.04-3.15) | 0.350 |
| Moxifloxacin addition | 4 (9.5) | 2 (22.2) | 2.71 (0.41-17.78) | 0.298 |
| Drug resistance | 4 (10.5) | 2 (22.2) | 2.43 (0.37-15.95) | 0.356 |
| Positive smear | 9 (21.4) | 2 (22.2) | 1.05 (0.18-5.94) | 0.958 |

IQR, interquartile range; ACKD, advanced chronic kidney disease; TB, tuberculosis. P-values were calculated using the Mann-Whitney U test for age and Fisher's exact test for categorical variables.

## DISCUSSION

Our matched analysis showed no excess one-year mortality in ACKD, with high cure and low relapse, despite widespread use of a hemodialysis-aligned dosing schedule necessitated by limited single-agent availability. These findings support pragmatic, dialysis-timed dosing in resource-limited settings, consistent with contemporary standards that recommend interval-extended dosing of renally cleared agents and careful management of rifampicin interactions.

Notably, there was a high rate of disseminated TB cases. This could be explained by the high proportion of immunosuppression, referral bias to a tertiary care center, and the fact that our institution is not the primary facility treating pulmonary TB in the region. TB incidence has been studied at our center from 2000 to 2015; 533 cases were evaluated, and 161 (30.2%) were caused by *M. bovis*. Characteristics associated with *M. bovis* disease were younger age, glucocorticoid use, and extrapulmonary disease (12).

Distinct features of extrapulmonary and disseminated tuberculosis in HIV- and non-HIV-associated immunosuppression were also previously evaluated in our center. *M. bovis* was responsible for 44% (80 out of 180) of the total tuberculosis cases in the study cohort, and they were significantly more common in the non-HIV-associated immunocompromised group, where they accounted for 54% of cases (44 of 81 patients) (13).

In Nepal, Pradhan et al. studied 49 CKD patients with TB, finding extrapulmonary disease in 69.1% and pulmonary disease in 21.8%. The most frequent extrapulmonary sites were the pleura, lymph nodes, and pericardium. Only 12.3% were microbiologically confirmed, with the rest (87.7%) treated empirically due to difficulties obtaining diagnostic samples (14). Our work stands out because most of our TB cases were pulmonary, and diagnoses were microbiologically confirmed in the majority.

In our analysis, ACKD did not increase one-year all-cause or TB-attributable mortality, and cure rates were comparable to those of the non-ACKD group, despite the use of the unvalidated alternating hemodialysis regimen employed at our institution. These findings contrast with reports of markedly worse outcomes in some settings. We found no significant differences in one-year all-cause mortality or TB-related mortality between ACKD and non-ACKD patients. Both groups had similar cure rates, with no documented relapses. Only the control group had TB-related deaths.

The only factors associated with increased mortality were rheumatologic disease and hepatotoxicity. The association with rheumatologic disease likely reflects the burden of severe immunosuppression. The strong link with hepatotoxicity is notable; while it may be a direct cause of morbidity, it could also be a marker of severe disease or underlying metabolic dysfunction.

International studies on CKD and TB outcomes show mixed results. In China, Xiao et al. studied 167 TB patients, finding higher mortality in ACKD (58% vs 6.1%) and higher treatment failure rates. Age >40, hypoalbuminemia, and CKD on HD were independently associated with mortality (15). Unlike our findings, their ACKD mortality was markedly higher. In Japan, Saito et al. found no differences in in-hospital TB mortality between ACKD and non-ACKD (3.4% vs. 7.1%) (16), which aligns with our results. Igari et al. analyzed 759 hospitalized TB patients (2007–2012) and found lower cure rates (52.7% vs 67.3%) and higher mortality rates (25.4% vs 12.4%) among patients with CKD. Hypoalbuminemia, glucocorticoid use, and cardiovascular disease were associated with worse outcomes (17). In India, Vikrant et al. studied 32 TB patients on RRT; 75% had extrapulmonary TB, 11 had peritoneal TB, and mortality reached 47% (60% in PD, 35.3% in HD), with peritoneal TB having the worst prognosis (18). In Turkey, Taskapan et al. followed 296 HD patients, of whom 6% developed TB. Mortality was higher in patients with delayed diagnosis and initiation of RRT (19). Our study did not analyze the time interval from RRT initiation to TB diagnosis, which represents an opportunity for future research. Differences across studies likely reflect variations in case mix (e.g., peritoneal TB burden), access to diagnostic tools, timing of RRT, and immunosuppressive comorbidities.

Several factors may explain this discrepancy. First, our center’s advanced diagnostic capabilities enabled a high microbiological confirmation rate (88%), potentially leading to earlier and more accurate diagnoses than in settings reliant on empirical treatment. Second, our control group had a high burden of immunosuppression (e.g., 21% HIV prevalence), which may have elevated their risk, narrowing the outcome gap with the ACKD group. Finally, the structured, multidisciplinary follow-up at a tertiary referral center likely played a crucial role in optimizing care for patients with complex ACKD.

When evaluating the reported adverse events (AEs), no significant differences were found between the two groups. Three patients with AEs were reported in the CKD group, only one of them being a case of hepatotoxicity, which led to the need for the temporary use of second-line parenteral medications. Hepatotoxicity was significantly associated with mortality in the overall cohort, although causality cannot be established.

In a Mexican study, Covarrubias-López et al. found >80% AE rate in 60 TB patients, mainly renal and hepatic dysfunction; one-third of those cases were in patients receiving second-line drugs (20). In the UK, Quantrill et al. reported AEs in 45.8% of CKD-TB patients — hepatotoxicity (4), gastrointestinal effects (4), and neuropsychiatric effects (6) — all in RRT patients (21). Vikrant et al. reported AEs in 29%, mostly gastrointestinal (12.9%) and hepatotoxicity (12.9%) (18). In China, Xiao et al. found higher rates of mild GI and neurotoxic AEs in ACKD patients (15).

In our center, the use of isoniazid or rifampicin for TB preventive treatment was evaluated in patients with type 2 diabetes. A total of 130 patients were assessed, with 68 randomized to isoniazid and 62 to rifampicin. The study was prematurely halted because of adverse events. The isoniazid group had significantly more permanent treatment interruptions due to grade 2 recurrent or grade 3 or 4 hepatotoxicity. In comparison, the RIF arm had more treatment interruptions due to grade 3 or 4 gastrointestinal intolerance. Among the possible causes, it was hypothesized that a condition that may have favored hepatotoxicity was the pre-existence of undiagnosed and aggravated metabolic dysfunction-associated steatotic liver disease (22).

There is limited evidence regarding the treatment of TB in patients on PD. Specific pharmacokinetic and dosing data are scarce for this population, and recommended doses for patients on HD may not be directly applicable to those on PD. These patients require close monitoring to detect toxicity; therefore, it is recommended that serum concentrations of anti-tuberculosis drugs be measured before and after each session. In the absence of definitive evidence, current guidelines recommend either using the doses for HD patients or switching renal replacement therapy (RRT) from PD to HD during TB treatment (8,9).

In our study, we analyzed four patients with PD. One of these patients died during the following year from a cause unrelated to TB, while the rest completed treatment without incidents, and no adverse events were reported in any of them. Further studies are needed in this population to determine the most effective treatment strategies, as our work does not permit drawing solid conclusions.

The primary challenge in treating TB in ACKD in Mexico is the lack of single-drug formulations of pyrazinamide and ethambutol. Our “alternating regimen” was developed as a pragmatic solution to align with hemodialysis schedules and approximate guideline-recommended, dose-adjusted therapy. The excellent outcomes observed suggest that with careful management, this approach can be practical. This is a critical finding for clinicians in similar resource-constrained environments across Latin America and beyond, where fixed-dose combinations are the norm. It demonstrates that effective TB treatment in ACKD is achievable even without ideal drug formulations, provided a structured, monitored approach is used.

This study provides real-world evidence on TB drug adjustment in patients with ACKD, including all eligible cases matched by age and sex, which strengthens the observed associations. It stands out for achieving a high microbiological confirmation rate, overcoming a standard limitation in extrapulmonary TB. At our Institution, all biopsies, cerebrospinal fluid samples, bronchoalveolar lavage, bone marrow specimens, and medical devices sent to the Microbiology Laboratory for culture are also cultured for mycobacteria, thereby increasing our diagnostic yield. Additionally, it offers a detailed analysis of ACKD etiologies and includes a control group with other comorbidities, thereby enhancing the validity of the observed association with mortality. Notably, this is the first formal description of an alternative hemodialysis-based TB treatment strategy in Mexico, demonstrating both safety and efficacy.

Our study has several limitations. Its retrospective design and modest sample size from a single tertiary center limit generalizability and preclude a multivariate analysis to adjust for potential confounders. The high rate of microbiological confirmation and tertiary care context may not reflect conditions in primary or secondary care settings. Furthermore, we did not perform therapeutic drug monitoring, which is the standard of care in many high-income settings (7). The small number of patients on peritoneal dialysis (n=4) precludes definitive conclusions about optimal management in this subgroup.

Implications. When individualized formulations are limited, a structured, dialysis-timed alternative regimen can be used without significant loss of effectiveness. Implementation research should evaluate standardized dialysis-aligned dosing, interaction management (especially with calcineurin/mTOR inhibitors), and the practicality of therapeutic drug monitoring in patients with ACKD and TB care.

### Conclusions

Among patients treated for TB at a Mexican tertiary center, ACKD was not associated with worse one-year mortality or cure rates compared to non-ACKD patients. A pragmatic, alternating hemodialysis-aligned regimen was associated with favorable outcomes and may represent a feasible and effective strategy in resource-limited settings where individual drug formulations are unavailable. These findings warrant validation in larger, prospective, multi-center studies.

## Data Availability

All relevant data are within the manuscript and its Supporting Information files.

